# Metabolomic analysis of plasma improves the prediction from hyperuricemia to gout incidence

**DOI:** 10.1101/2025.09.22.25336391

**Authors:** Zhao-Hui Ruan, Jie Zhang, Si-Qin Xia, Hua-Hui Zhu, Cheng Zhou, Zi-Xing Zhang, Dong-Qing Ye, Xin-Yu Fang

**Affiliations:** Department of Epidemiology and Biostatistics, School of Public Health, Anhui Medical University, Hefei, Anhui 230032, China; Inflammation and Immune Mediated Diseases Laboratory of Anhui Province, Hefei, Anhui 230032, China; School of Public Health, Anhui University of Science and Technology, Hefei, Anhui 231131, China ZH-R and JZ contributed equally

**Keywords:** Metabolomics, gout, hyperuricemia, prediction

## Abstract

**Objectives:** To investigate the metabolome perturbation in the progression of hyperuricemia (HUA) into gout, and evaluate the predictive power of metabolomics.

**Methods:** Circulating metabolomics data from 24225 individuals were measured using nuclear magnetic resonance (NMR) technology, and we used the Cox models to assess the hazard ratios of metabolites in HUA-to-gout progression. Key metabolites were selected through 10-fold cross-validated elastic net regression, with 10-year prediction models developed using multivariate Cox regression. The predictive performance was differentiated by comparing the area under the receiver operating characteristic curves (AUCs). We used Net Reclassification Improvement (NRI) to estimate the improvement in reclassification ability with the addition of metabolites to the conventional prediction model.

**Results:** Of the 24225 HUA patients, the median follow-up period was 13.6 years, during which 1584 participants developed gout. 18 metabolites showed significant associations; the most positive association was with glycoprotein acetyl (HR 1.10; 95% CI: 1.04, 1.16) and the most negative association was with IDL particle concentration (HR 0.91; 95% CI: 0.87, 0.96). The predictive ability (AUC: 0.80 vs 0.78) and reclassification ability (NRI = 2.83%, *P* < 0.001) of the new combined model were significantly improved with the addition of selected metabolites (n = 44), allowing the identification of high-risk groups.

**Conclusions:** Our analyses identified various metabolomic profiles significantly associated with the development of HUA into an incident event of gout, and implied that metabolomics can enhance predictive accuracy for clinical progression from HUA to gout.

## Introduction

Gout is a type of swelling and damage in the body caused by the build-up of sodium urate in bones, joints, kidneys[1, 2], and under the skin. It is linked to high levels of uric acid in the body due to problems with how the body processes purines and/or how it gets rid of uric acid. This affects about 6. 8% of people worldwide[3]. Gout is chronic and recurrent, often accompanied by intolerable pain, and with the accumulation of time the joints will be gradually destroyed and gout stones will appear, which will lead to a variety of deformities and disabilities, renal impairment, and even lead to heart failure, ischaemic stroke and death.[4]. Research has found that people with gout have to pay a lot of money for healthcare, from $172 to $6179[5], for going to the emergency room a lot, staying in the hospital often, not being able to do as much at work, and having trouble doing regular activities [6–8]. In short, gout can be very expensive and stressful for patients, and it can really affect their quality of life and how long they live in good health. HUA is a very important process in causing gout. Past research has focused on the connection between HUA and gout[9–11]. Gout happens in only one out of three people with HUA [12]. HUA has a high prevalence and therefore comprehensive intervention is difficult to achieve. Identifying people with HUA who are at risk of having a gouty attack and targeting them for early intervention can reduce the risk of gout onset, which is an important clinical practice issue and a public health issue worthy of research.[13].

Metabolomics studies how metabolites change in the body when external stimuli affect it. It does this by discovering, identifying and measuring low molecular weight substances in blood samples[14, 15]. There aren’t many studies on how high uric acid levels can lead to gout, most studies have looked at how metabolites in the body are different, but not many have looked at how these metabolites can predict who will get gout[16, 17]. Some studies have developed multivariate models based on the Random Forest unbiased variable selection algorithm to differentiate between patients with HUA and gout, but the sample sizes of the studies were small and the predictive models developed included few metabolite species[10]. In addition, there was a study that used multivariate selection algorithms to screen metabolites and developed predictive models to predict whether a given patient would have a rare attack of gout or a frequent attack of gout, but this was a single-center research which had a limited number of participants and the study did not involve patients with HUA[18].

In this study, we used data from the UK Biobank, measured using a standardized platform of nuclear magnetic resonance (NMR) technology, showing changes in chemicals in the human body over time. These data come from large samples of people who have been followed up over a long period of time. We used these data to screen for metabolites that play a role in the progression of HUA to gout, and developed traditional risk factor prediction models, metabolite prediction models, and combined prediction models combining metabolites and traditional risk factors to differentiate between patients with HUA and those with gout, respectively and to predict the progression of the disease.

## Materials and Methods

### Study population and data collection

The UK Biobank is a study sponsored by the Medical Research Council and the Wellcome Trust. More than 500,000 people aged 40-69 years in the UK participated in the study between 2006 and 2010, completing touch-screen questionnaires at each of the 22 assessment centers under the guidance of a professional staff member, and obtaining biospecimens and body measurements matched to health-related records. Previous articles have provided detailed information on the UKB study design and participants. The study was conducted under the approval of the North West Multi-center Research Ethical Committee (11/NW/ 0382). All participants provided written informed consent.[19].

To identify metabolomic biomarkers associated with the progression of HUA to gout, patients with HUA who did not have gout and for whom metabolite data were available were included in this analysis. Of the initial 502359 participants, 7960 with gout were first excluded, followed by 237445 and 11523 participants excluded because of missing metabolite and uric acid data respectively, followed by 206552 participants excluded according to the inclusion criteria for HUA, and finally outliers in the data and missing data for the variables, resulting in a final total of 24225 participants who were enrolled in the study at baseline.

To develop a predictive model of the risk of developing gout in patients with HUA over 10 years, participants were assigned to the training set (n = 19380) and validation set (n = 4845) in a 4:1 ratio randomly. Figure S1 shows the flowchart of this study. Baseline characteristics of the observed subjects in the overall and each dataset are described, respectively in Table 1 and Table S1.

**Table 1.**
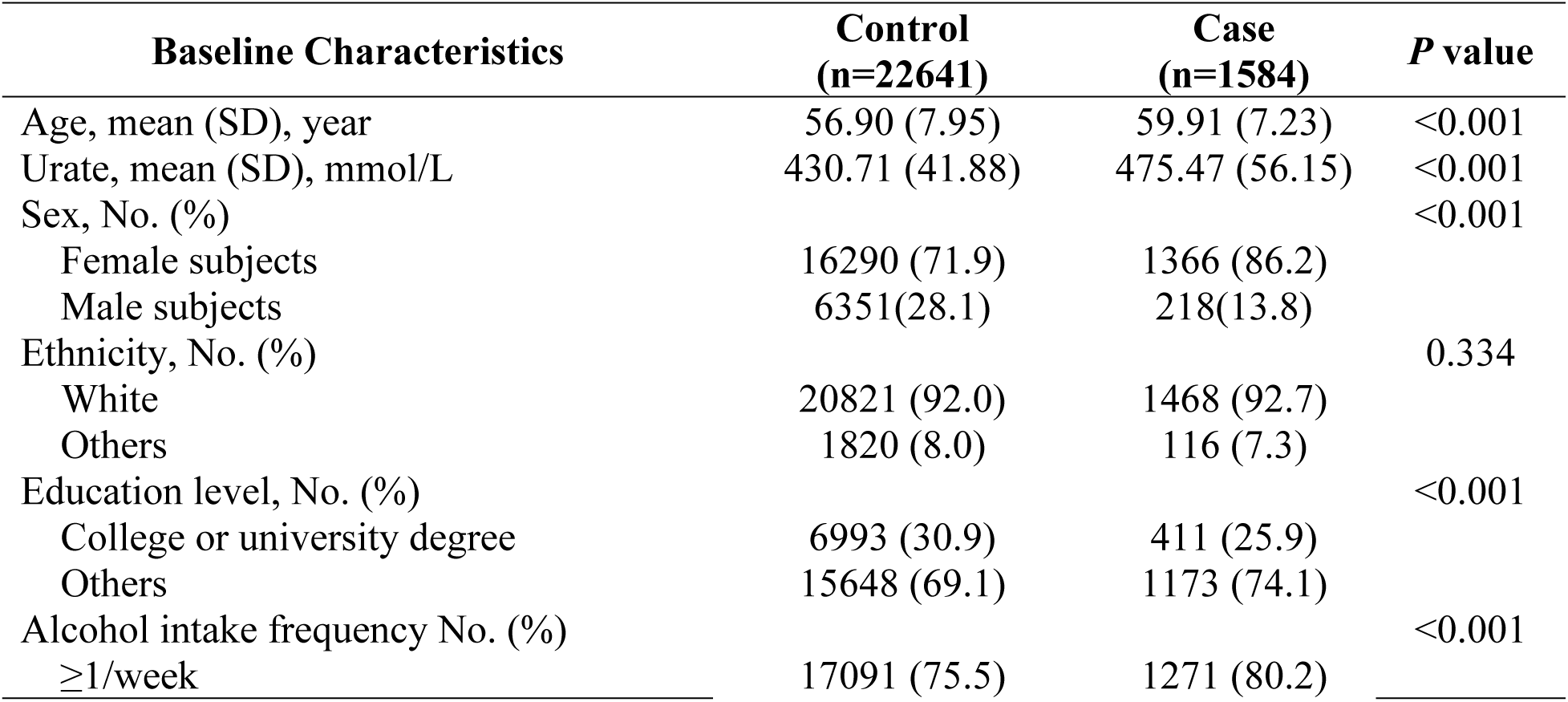
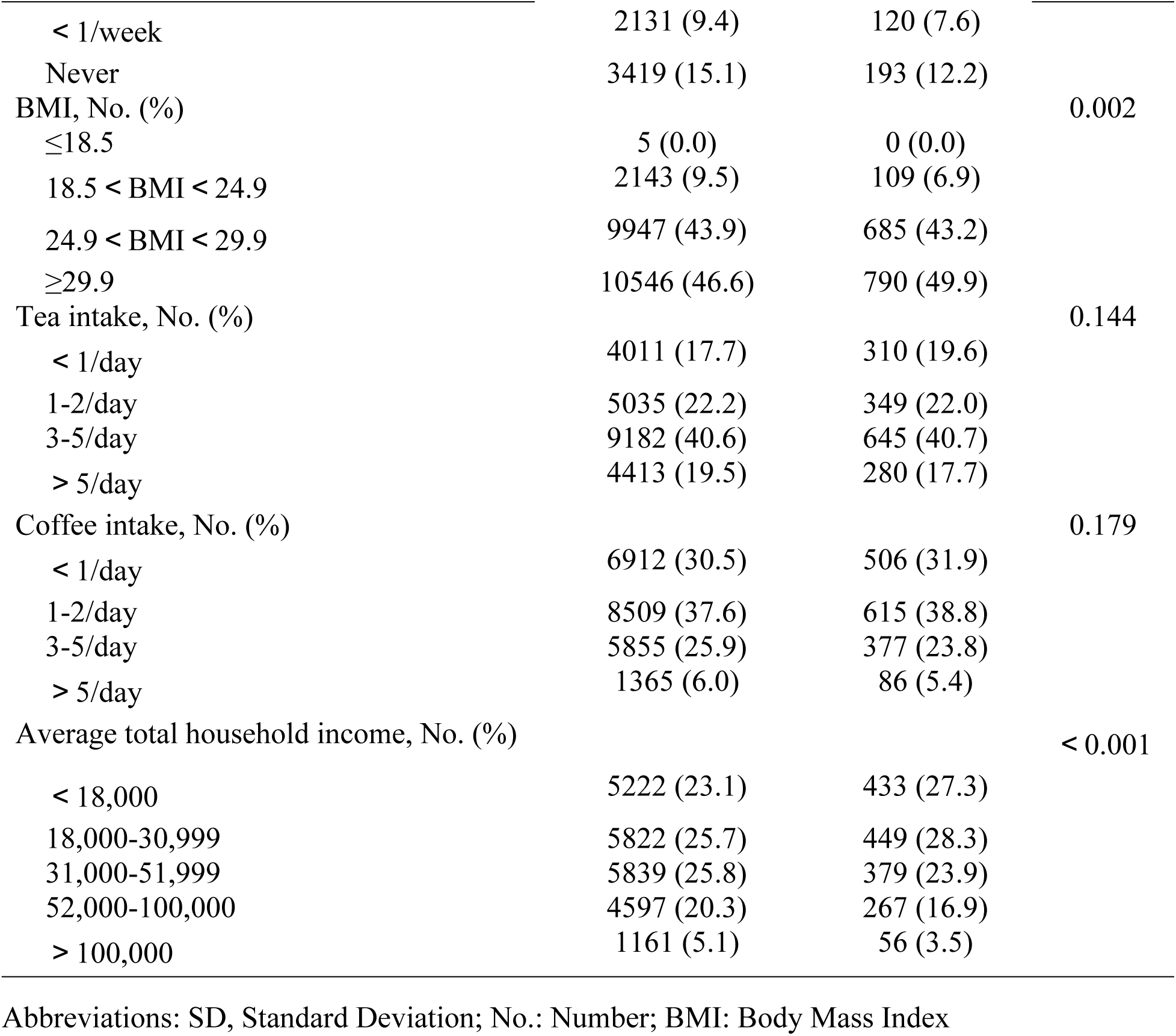
Baseline characteristics of all study participants in the UK Biobank.

| Baseline Characteristics | Control<br>(n=22641) | Case<br>(n=1584) | *P* value |
| --- | --- | --- | --- |
| Age, mean (SD), year | 56.90 (7.95) | 59.91 (7.23) | <0.001 |
| Urate, mean (SD), mmol/L | 430.71 (41.88) | 475.47 (56.15) | <0.001 |
| Sex, No. (%) |  |  | <0.001 |
| Female subjects | 16290 (71.9) | 1366 (86.2) |  |
| Male subjects | 6351(28.1) | 218(13.8) |  |
| Ethnicity, No. (%) |  |  | 0.334 |
| White | 20821 (92.0) | 1468 (92.7) |  |
| Others | 1820 (8.0) | 116 (7.3) |  |
| Education level, No. (%) |  |  | <0.001 |
| College or university degree | 6993 (30.9) | 411 (25.9) |  |
| Others | 15648 (69.1) | 1173 (74.1) |  |
| Alcohol intake frequency No. (%) |  |  | <0.001 |
| ≥1/week | 17091 (75.5) | 1271 (80.2) |  |
| < 1/week | 2131 (9.4) | 120 (7.6) |  |
| Never | 3419 (15.1) | 193 (12.2) |  |
| BMI, No. (%) |  |  | 0.002 |
| ≤18.5 | 5 (0.0) | 0 (0.0) |  |
| 18.5 < BMI < 24.9 | 2143 (9.5) | 109 (6.9) |  |
| 24.9 < BMI < 29.9 | 9947 (43.9) | 685 (43.2) |  |
| ≥29.9 | 10546 (46.6) | 790 (49.9) |  |
| Tea intake, No. (%) |  |  | 0.144 |
| < 1/day | 4011 (17.7) | 310 (19.6) |  |
| 1-2/day | 5035 (22.2) | 349 (22.0) |  |
| 3-5/day | 9182 (40.6) | 645 (40.7) |  |
| > 5/day | 4413 (19.5) | 280 (17.7) |  |
| Coffee intake, No. (%) |  |  | 0.179 |
| < 1/day | 6912 (30.5) | 506 (31.9) |  |
| 1-2/day | 8509 (37.6) | 615 (38.8) |  |
| 3-5/day | 5855 (25.9) | 377 (23.8) |  |
| > 5/day | 1365 (6.0) | 86 (5.4) |  |
| Average total household income, No. (%) |  |  | < 0.001 |
| < 18,000 | 5222 (23.1) | 433 (27.3) |  |
| 18,000-30,999 | 5822 (25.7) | 449 (28.3) |  |
| 31,000-51,999 | 5839 (25.8) | 379 (23.9) |  |
| 52,000-100,000 | 4597 (20.3) | 267 (16.9) |  |
| > 100,000 | 1161 (5.1) | 56 (3.5) |  |

Abbreviations: SD, Standard Deviation; No.: Number; BMI: Body Mass Index

### Metabolic biomarker quantification

Blood samples from most of the participants in the UKB were tested via the MRI metabolic biomarker platform between 2006 and 2010, and a total of 249 metabolic markers were measured. These included 168 levels and 81 ratios to study their metabolism in relation to lipoprotein lipids, fatty acids and their composition, and a variety of other low molecular weight metabolites, and the detailed implementation of the MRI experiments has been described in other articles[20–22]. In our study, the focus was on those metabolites that could be measured directly and those that could not be inferred, and a total of 143 metabolites were selected for analysis[23].

### Ascertainment of gout and HUA

Participants who met any of the following criteria were recognized as gout patients [24, 25]: self-reported physician-diagnosed gout; a prescription for uric acid-lowering therapy (ULT) without a clinically confirmed diagnosis of lymphoma or leukemia (International Classification of Diseases (ICD)-10 codes C81-C96); and discharge records with a primary or secondary diagnosis of gout (ICD-10 codes: M10, M100-M14 and M109).

Among participants without gout, the level of serum urate ≥400 µmol/L for males or ≥360 µmol/L for females was considered as HUA[26].

### Ascertainment of risk factors

Risk factors were identified from data collected from the UK Biobank baseline assessment and relevant medical records. age[27], sex[28], body mass index[29], ethnicity[3], alcohol intake frequency[30], urate level[31], qualifications[32], tea intake[33], coffee intake[33] and household income[34] were established risk factors for gout and are therefore used as covariates in Cox regression analyses and constitute conventional predictive models for the progression of HUA to gout.

Age and urate level were continuous variables, sex, ethnicity, body mass index (less than or equal to 18.5/18.5 to 24.9/24.9 to 29.9 or greater than or equal to 29.9 kg/m2), alcohol intake frequency (greater than or equal to once a week/less than once a week or never), qualifications (college or university degree or others), tea intake (less than 1 cup per day/1 to 2 cups per day/3 to 5 cups per day or greater than 5 cups per day), coffee intake (less than 1 cup per day/1 to 2 cups per day/3 to 5 cups per day or greater than 5 cups per day), household income (less than 18000/18000 to 30999/31000 to 51999/52000 to 100000 or greater than 100000) were redefined as categorical variables.

### Statistical analyses

Means (standard deviation, SD) or medians (interquartile range, IQR) were used to describe continuous variables, and numbers and percentages were used to describe categorical variables. For all metabolite values, a natural logarithmic transformation (ln[x+1]) followed by a Z transformation was performed, and metabolite levels exceeding a median of 4 IQR were considered outliers and excluded.

Our study consisted of two different analyses, and the flow chart is shown in Figure 1. In the first analysis, a Cox proportional hazards model was used to estimate the relationship between the progression of HUA to gout and metabolites, with confounders chosen to be age, sex, body mass index, ethnicity, frequency of alcohol intake, uric acid levels, educational qualifications, tea intake, coffee intake and household income. The false discovery rate[35] (FDR)-corrected threshold was set to p<0.05. Table S2 and Figure S1 show the FDR P-values, hazard ratios (HR) and 95% coefficient intervals (CI) for all 143 metabolites in the Cox model.

**Figure 1.**
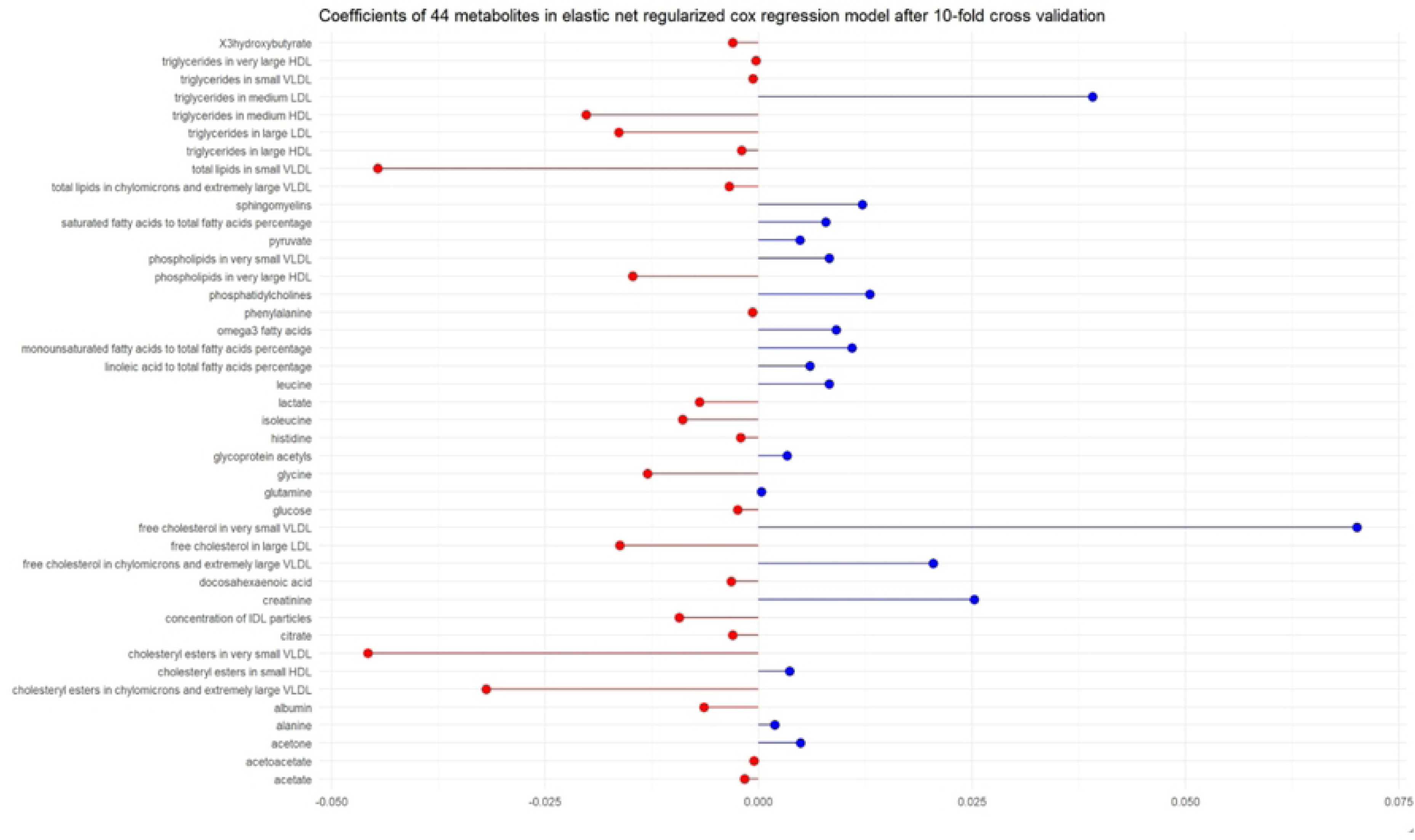
Lollipop chart of selected 44 metabolites in an elastic net regularized Cox regression model after 10-fold cross-validation.

The second analysis was conducted in three steps to develop and validate a risk prediction model for the progression of HUA to gout over 10 years. We divided the participants into two groups, a training set and a test set. Then, we used a special math model to pick out the most important metabolomic predictors. The elastic net regularized Cox regression model is a type of machine learning algorithm used for predicting diseases. It combines two methods called LASSO and Ridge regression to choose the best variables for prediction. The LASSO method helps to pick the most important variables by making the weights smaller, while the Ridge method helps to make the weights less extreme. This model is good at explaining the likelihood of disease and finding the right variables for prediction. A lot of information about the model can be seen in other articles[36]. The resilient net regression model uses two tuning numbers. One shows the weight of the penalty (α), and the other shows how complicated the penalty is (λ). It’s important to know that when α is 1, LASSO is preferred, and when α is 0, Ridge regression is preferred[37]. To achieve model sparsity and select core metabolome predictors, we tested different values of α (0.5, 075 and 1). We then used a 10-fold cross-validation approach to select the best values of λ and β in the elastic net regression model to achieve the best robustness of the model[38]. We performed 10-fold cross-validation on a 2D grid for different combinations of α and λ to calculate the cross-validation prediction error for the elastic net Cox regression in order to achieve the smallest cross-validation function and to assign β coefficients to each metabolite to gain the smallest cross-validation function, and finally selected α = 1 and λ = 0.0004259433, which resulted in the smallest cross-validated mean deviation of 0.06026365. Table 2 lists the 44 metabolites with non-zero coefficients. In the second step, we constructed three predictive models in the validation set and used Cox regression analysis to estimate the predictive power of the different models separately. Model 1 included traditional risk factors, including age, gender, body mass index, ethnicity, frequency of alcohol intake, uric acid level, seniority, tea intake, coffee intake, and household income; model 2 included selected metabolomic biomarkers; and model 3 included a combination of traditional risk factors and selected metabolomic biomarkers. The coefficients (95% CI) and P values for each exposure parameter are shown in Table S3. In the final step, the predictive value of the 44 selected metabolites was assessed by two methods. First, receiver operating characteristic curves (ROCs) were built to compare the area under the curve (AUC) of the different models. We then estimated the extent to which the screened metabolomic biomarkers contributed to risk stratification for the development of gout compared with traditional risk factors by calculating a net reclassification index (NRI), and if the NRI exceeded 5%, we considered these HUA patients to be at high risk of developing gout in the next 10 years.

**Table 2.** Net reclassification improvement of adding 44 metabolites to a conventional risk prediction model.

|  |  | Conventional<br>prediction model | Updated model |
| --- | --- | --- | --- |
| Incidence of gout in 10<br>years | Risks(%) | < 5 | ≥ 5 |
| YES | < 5 | 305 | 51 |
|  | ≥ 5 | 25 | 482 |
| NO | < 5 | 17569 | 663 |
|  | ≥ 5 | 628 | 3180 |

Conventional prediction model: risk factors model; updated model: combined model.

In addition, it has been shown that 540 mmol/L is usually the uric acid level at which treatment is effective[39] or relevant clinical symptoms appear[40] in gout patients, so we stratified the validation set population using 540 mmol/L as the threshold. They were divided into the high-urate group with uric acid levels higher than or equal to 540 mmol/L (n=139) and the low-urate group with uric acid levels lower than 540 mmol/L (n=4704), and ROC curves were constructed for each group to investigate the differences in disease prediction ability of the three different models at different uric acid stratification levels.

All analyses were performed using R (version 4.4.0, R Project for Statistical Computing, Vienna, Austria).

## Results

### Baseline characteristics of the study participants

A total of 24225 patients with HUA who did not have gout and who had completed NMR metabolomics measurements data were included in this study; these participants were free of missing data, had a mean age (standard deviation) of 57.1 (7.9) years, and were mostly female subjects (71.9%). The median follow-up was 13.6 years (IQR: 12.7-14.4 years), and 1,584 participants developed gout during the follow-up period. Compared to participants with non-gout, those who developed gout were more like female, older, overweight or obese, people with high uric acid levels, less educated, alcoholics, people with high tea and coffee intake and low-income families. Table 1 shows the baseline characteristics of all participants (stratified by gout onset).

### Associations of baseline Circulating Metabolites and progression of HUA to gout

After adjusting for age, sex, body mass index, ethnicity, frequency of alcohol intake, uric acid level, seniority, tea intake, coffee intake, and household income, a total of 18 of the 143 metabolic biomarkers showed significant associations with the progression of HUA to gout in the Cox models at FDR-controlled *P*<0.05. These metabolites included cholesterol, esterified cholesterol, fatty acids, free cholesterol, ketone bodies, fluid balance and glycoprotein acetyls, lipoprotein particle, phospholipids, size and apo-LP and total lipids. Of the 18 metabolites, only glycoprotein acetyls seemed to be a risk factor for the progression of HUA to gout (HR=1.0992, 95% CI 1.043 to 1.158), inverse with the remaining 17 metabolites. (Figure S2)

### Metabolite selection, prediction and reclassification of the likelihood of progression of HUA to gout

Elastic net regularized Cox regression analysis identified metabolites for use in constructing a predictive model for progression of HUA to gout, and 44 metabolites were selected for inclusion in the training set (Table S4 and Figure 1). The AUC of the traditional risk factor-based prediction model (risk factors model) was 0.78, and the AUC of the selected metabolites model (selected metabolites model) was 0.70, which shows that the accuracy of the above two models in predicting the onset of gout is limited. However, the AUC of the combined model, which combines these metabolites with traditional risk factors, increased to 0.80, and the predictive accuracy of the new model was significantly improved (Figure 2). After adding metabolic predictors to the traditional prediction model, the net reclassification ability of the new model was significantly improved. (NRI=2.83%, SE=0.002, *P*<0.001) (Table 2).

**Figure 2.**
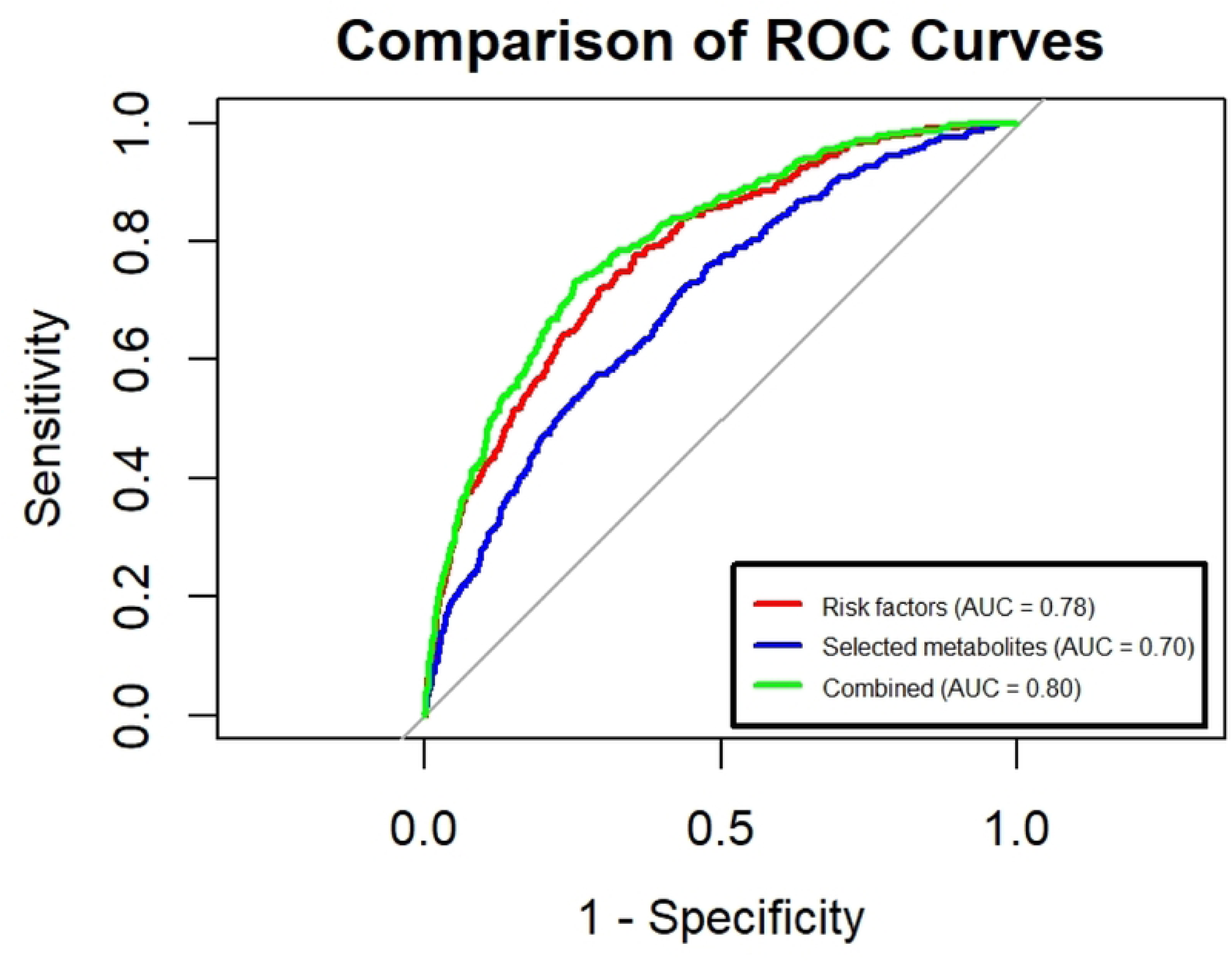
ROC curves for the three models.

In the uric acid stratification analysis, the predictive power of metabolites in the low-urate group remained poorer than in the conventional model (AUC: 0.69 vs. 0.76). The addition of metabolites to the conventional model also improved the predictive ability (AUC: 0.76 vs. 0.79). However, in the high-urate group, the predictive ability of metabolites was better than that of the traditional model (AUC: 0.81 vs. 0.62), and the predictive performance of the combined model was significantly improved by adding metabolites to the traditional model (AUC: 0.62 vs. 0.83) (Figure 3 and Figure 4). The results of the stratified analyses revealed that the predictive performance of the three models in the low-urate group with uric acid levels below 540 mmol/L was consistent with the findings of the total population. However, the opposite findings were seen in the high-urate group with uric acid levels higher than or equal to 540 mmol/L, where, unlike the total population, the predictive power of the metabolites was significantly higher and higher than that of the traditional risk factors. In addition, the predictive performance of the combined model was also higher in the high-urate group than in the total population and the low-urate group, suggesting that the predictive ability of the metabolites increased with the uric acid level in this study, and the predictive ability of the new model increased as well.

**Figure 3.**
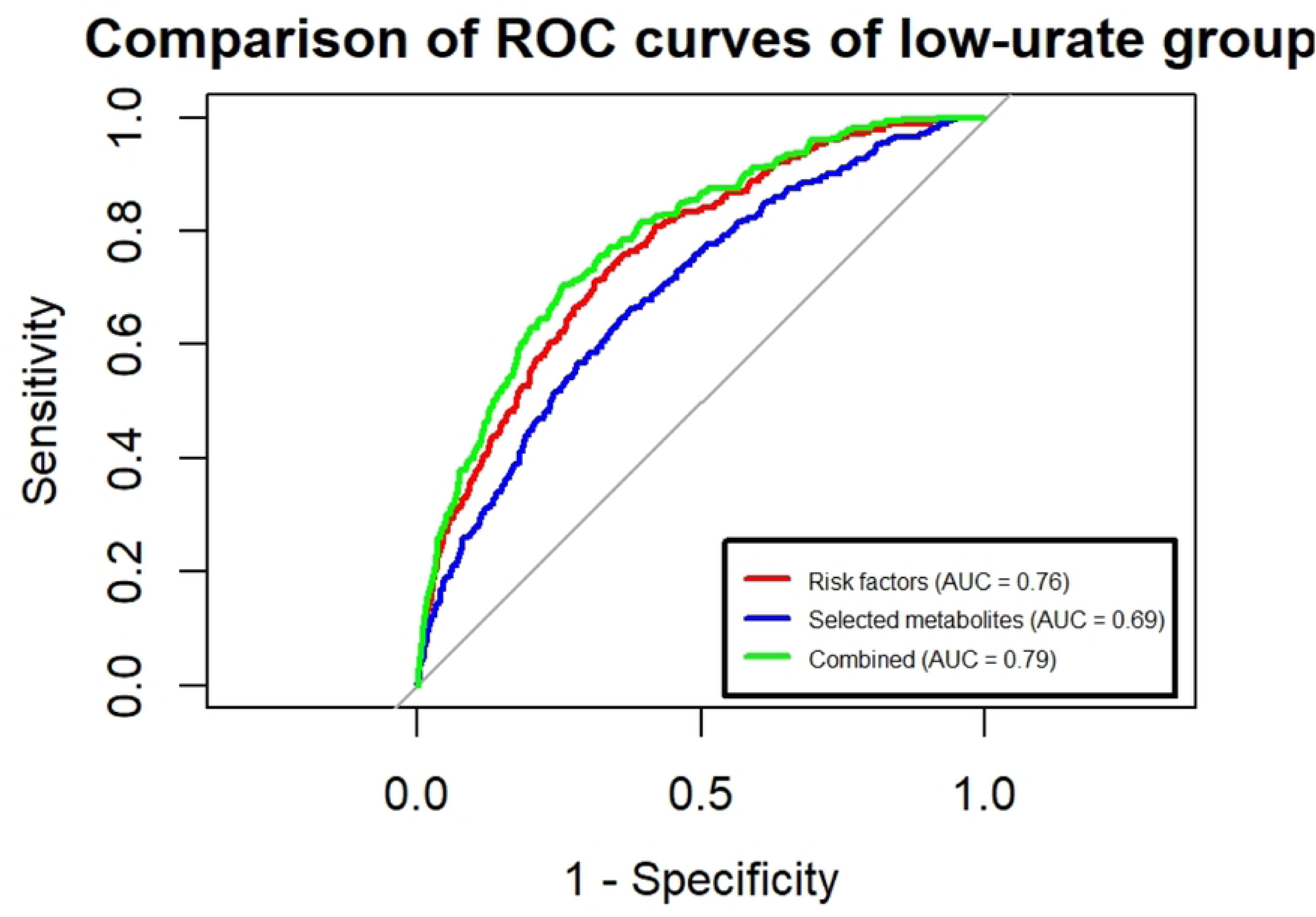
Comparison of ROC curves of the low-urate group.

**Figure 4.**
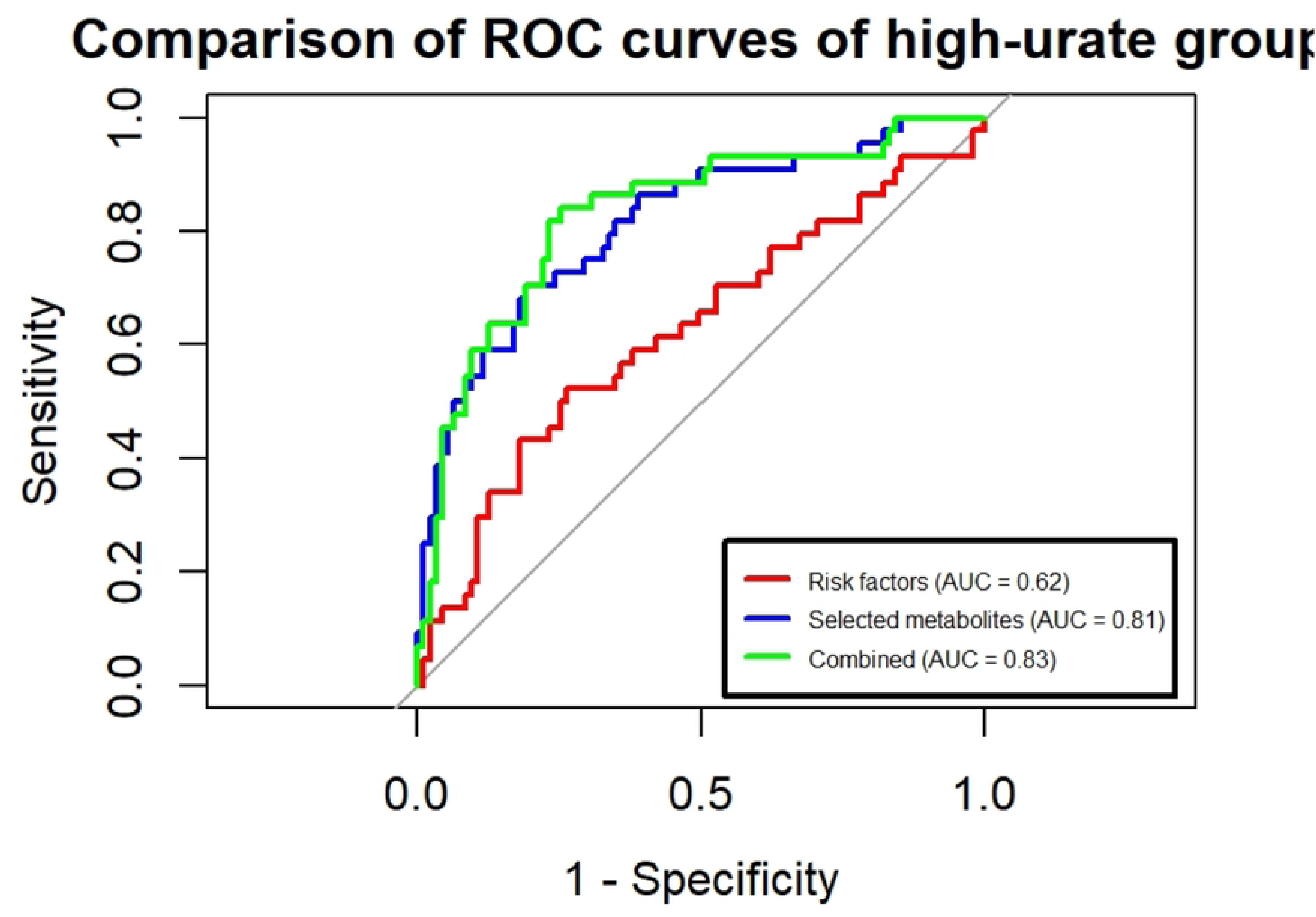
Comparison of ROC curves of the high-urate group.

## Discussion

Using data from over 24225 samples from the UK Biobank, we conducted a large sample size metabolomics study to identify metabolic profiles associated with the gouty process in patients with HUA, screening for 28 relevant metabolites including cholesterol, phospholipids, and glycoprotein acetyls, among other materials. More importantly, we screened 44 metabolite predictors using the elastic net regularized Cox regression model and constructed a conventional model containing only conventional risk factors (risk factors model), a predictive model containing only metabolites (selected metabolites model), and a combinatorial model consisting of a combination of the two (combined model), with the AUCs were 0.78, 0.70 and 0.80. Good sensitivity and specificity were achieved in predicting HUA patients with the potential to develop gout using the three models described above. The results of this study suggest that glycoprotein acetyls (GlycA) is a risk factor in the progression of HUA to gout, which is consistent with other studies[41]. It has been shown that the association of GlycA with the onset and recurrence of gout increases progressively with fasting time[41]. GlycA is a tool that shows the level of inflammation in the body. It measures the levels of certain proteins, such as cryptococcal protein and alpha-1-acid glycoprotein, that the body releases in response to infection or disease[42]. It is believed that GlycA levels in the body are related to how genes work together in neutrophils, a type of white blood cell that helps fight infection. This relationship involves many genes that are important for maintaining the function of neutrophils. Neutrophilic synovitis, which is a key part of gout attacks, happens when the body’s natural defense system fights against deposits of a substance called monosodium urate crystals. This causes inflammation in the joints. The body’s response to the crystals activates a process that releases molecules that cause more inflammation and attract more immune cells to the area[43]. The present study supports the risk of GlycA in the development of gout, and the initiation of further studies could help to gain insight into the metabolic-neutrophilic synovitis pathway in gout and have implications for biomarker-based prediction of gouty attacks or therapeutic targeting.

In the present analysis, lipid composition (including cholesterol, esterified cholesterol, free cholesterol, total lipids, fatty acids, and phospholipids) is observed to be a protective factor for the progression of HUA to gout, which is inconsistent with previous studies[44–46]. Furthermore, previous findings suggest that apolipoprotein B (apo B) is a risk factor for gout, but the observation in the present study that apo B is a protective factor for the progression of HUA to the gout process is not necessarily inconsistent with extant experimental results. Evidence from a study[47] suggests that a decrease in total apo B is associated with HUA compared to HUA, whereas an increase in total apo B is associated with gout, thus suggesting that Apo B has multiple effects on the development of gout.

More importantly, we identified 44 candidate metabolites through elastic net regularized Cox regression models, the addition of which to conventional models with traditional risk factors could significantly improve the prediction of the progression of HUA to gout and the accuracy of reclassification of different risk groups for the development of gout, which could improve the identification of patients at risk of disease progression and the delivery of targeted interventions, which suggests that metabolomic biomarkers have potential clinical use as population-based early predictors of gout disease progression.

In addition, by stratifying uric acid levels, the predictive model showed a similar trend in the HUA population with lower uric acid levels as the overall population, with a higher predictive ability for risk factors than for metabolites. However, in the HUA population with high uric acid levels, models containing metabolites have much stronger predictive power than traditional models containing risk factors, and the predictive power of combined models in this population is higher than that in the training set population. Therefore, it can be inferred that metabolites play a more significant role in the development of HUA to gout than risk factors in the HUA population with higher levels of uric acid. However, in the training set of this study, only 139 individuals had uric acid levels higher than 540mmol/L. Due to the small sample size, this conclusion needs to be further replicated in a larger sample to have stronger persuasiveness.

Our study has several strengths, firstly, it used data with a larger sample size compared to other studies in the same direction and the participants had a follow-up period of more than 10 years, which enhances the quality of the study, secondly, it used the homogeneous platform of metabolites analyzed using NMR technology, which ensures the accuracy of the data analysis, and lastly, our study combines not only metabolomics and machine learning algorithms, but also metabolites and conventional risk factors, and builds a prediction model that includes different factors, trying to identify the risk of gout development in HUA patients from diverse perspectives, which provides a theoretical basis for disease prediction.

There are also some limitations of this study: firstly, the majority of participants were white British people of good socio-economic status, which may have led to selection bias, and therefore the findings are worthy of replication for people with HUA and gout of other countries or ethnicities; secondly, the metabolites used in the study were limited in type and insufficiently broad in coverage, besides, metabolomics measurements were only performed during the baseline period, and may fluctuate during follow-up, and the key window of action is unknown; thirdly, complex drug and dietary interactions are not taken into consideration, which would have affected the metabolic profile of the participants; and finally, when stratifying uric acid levels, the number of people in the high-urate group of the training set was relatively small, and more in-depth studies with larger sample sizes are needed.

## Conclusions

Our results show that the progression of HUA to gout is significantly associated with metabolomic profiles, and that the new prediction model combining metabolites with traditional risk factors has good predictive performance, suggesting that metabolomics can improve the prediction of risk for clinical gout development.

## Acknowledgments

The authors would like to thank the UK Biobank participants. This study has been conducted using the UK Biobank resource under application number 62663.

## Ethics statements

This study involves human participants and was approved by the North West Multi-Centre Research Ethics Committee (11/NW/0382). Participants gave informed consent to participate in the study before taking part.

## Data availability statement

UK Biobank data can be requested from the UK Biobank. Data used and/or analyzed during the study are available from the corresponding author upon reasonable request.

## Supporting information

Figure S1. Flowchart of study participants in the UK Biobank.

Table S1. Baseline characteristics of the observed objects in the training and validation sets.

Table S2. β coefficient of adjusted HR (95% CI) and *P* values of Incident gout for 143 Metabolites in Cox Proportional Hazards Model.

Table S3. The coefficients (95% CI) and P-values of each exposure parameter for different models.

Table S4. Coefficients of Selected 44 metabolites in elastic net regularized cox regression model after 10-fold cross validation, their coefficients in e-net regression model in training dataset, and their VIFs in testing dataset.

Fig. S2. Volcano plot showing FDR *P*-values, hazard ratios (HR) and 95% coefficient intervals (CI) for 143 metabolites in the Cox model

